# Assessing the impact of social prescribing on wellbeing outcomes: A national analysis of UK data

**DOI:** 10.1101/2025.09.25.25336621

**Authors:** F. Bu, D. Hayes, L. Munford, D. Fancourt

**Affiliations:** Social Biobehavioural Group, Research Department of Behavioural Science and Health, Institute of Epidemiology & Health Care, University College London, 1-19 Torrington Place, London, WC1E 7HB; School of Health Sciences, University of Manchester, UK and NIHR Applied Research Collaboration Greater Manchester (ARC-GM), Manchester, UK

## Abstract

Social prescribing (SP) is growing rapidly in the United Kingdom and across the world. However, there remains a current lack of robust quantitative evidence on its impacts. Drawing pre-post data from large-scale longitudinal administrative records, this study aimed to provide the first large-scale national analysis of the wellbeing impacts of SP in the UK. Wellbeing outcomes were measured using the short Warwick-Edinburgh Mental Wellbeing Scale (SWEMWBS) and the Office for National Statistics wellbeing scale (ONS4), including happiness, anxiety, life satisfaction and worthwhile. Data were analysed using Bayesian growth curve modelling. Our findings demonstrate consistent and sizeable improvements across different wellbeing domains in the 1-6 months following initial referral to SP. Specifically, we observed a 3.31-point increase in SWEMWBS (95% HDI: 3.26-3.37), a 1.59-point increase in happiness (95% HDI: 1.55-1.63), a 1.57-point increase in life satisfaction (95% HDI: 1.54-1.61), a 1.4-point increase in life being worthwhile (95% HDI: 1.36-1.43), and a 1.45-point decrease in anxiety (95% HDI: -1.49 to -1.41). The increase in life satisfaction is equivalent to a conservative monetary estimate of £4,252 over a mean period of 2.5 months (2019 prices), representing a return on investment of £9 (per £1 invested). These results proved robust across multiple sensitivity analyses. We found little evidence that the wellbeing changes differ across socio-demographic groups, indicating a broad applicability of SP interventions.

## Introduction

Social prescribing (SP) is a mechanism of care which connects individuals to non-medical forms of support, based in their local communities [1]. In the last decade, SP has rapidly developed globally to help tackle social or complex health needs, with England leading the first nation-wide programme [1,2]. The most common model in England is the General Practitioner (GP) Link Worker (LW) model, in which LWs are employed through the Additional Roles Reimbursement Scheme (ARRS) to work directly with referred patients to co-develop a non-clinical plan based on patients’ values, needs and preferences, connecting them with existing community activities. These activities can include (but are not limited to) the arts, music, access to nature, volunteering, gardening, exercise, and broader support services [3]. In addition to the predominant GP LW model, individuals can also be referred via alternative routes, such as secondary care, voluntary and community sector organisations, statutory services, and self-referral. The importance of SP lies in its potential to address the social, emotional, and practical needs of patients that are often interrelated to medical needs but are not covered by clinical treatments, and which form an estimated 20% of GP consultations [4]. When incorporated into healthcare, SP can serve as a preventative approach, addressing challenges, such as social isolation and loneliness, either before physical and mental health symptoms arise or before they progress. Alternatively, it can be embedded into a patient’s healthcare plan alongside other treatments, to provide a holistic model of care [2].

To date, the evidence base for SP suggests an overall positive impact on wellbeing outcomes. A recent qualitative meta-synthesis across 18 studies of 1506 patients who received SP reported that SP increased wellbeing, via aspects such as increasing a sense of belonging to the community, increasing confidence and self-worth, and providing a sense of purpose, pride and achievement [5]. Quantitative metrics also suggest increases in wellbeing, with a recent systematic review identifying five studies where wellbeing was measured, of which four showed large effect sizes over a short follow-up period, whilst the other showed no effect over an intermediate follow up period [6]. However, whilst helpful, these reviews are not without limitations. The qualitative meta-synthesis included studies of varying quality, with participants being predominantly older adults and female, leaving it unclear whether other populations perceive similar benefits. Similarly, for the quantitative systematic review, studies also ranged in methodological quality, with most scoring weak ratings overall on quality assessment. Moreover, three studies [7–9] had sample sizes under 150 participants for pre-post outcome analyses, suggesting that they were likely to be underpowered to detect an effect. Additionally, as most studies focused exclusively on specific services or local areas, their findings cannot be generalised to broader services or patient populations. Lastly, the largest study (n=1297) examined only arts referrals via a local primary care setting [10], leaving an evidence gap on how SP referrals via non-primary care routes or to other non-arts services affect wellbeing.

Clearly, a key challenge in SP research is balancing different study designs. In light of the methodological limitations identified with previous studies outlined above, a number of controlled and randomised trials on SP are now underway [11,12]. While these will be critical for confirming causality, it is also important that national services are evaluated using large-scale administrative data that can capture real-world heterogeneity in service delivery and provide large samples enabling consideration of potential moderation by socio-demographic characteristics. In addition, given the current financial pressures experienced by the UK and other governments, it is particularly important to consider the opportunity costs and value for money of SP to support commissioning decisions.

In the UK, HM Treasury recommends that policies and interventions – such as SP LWs – are appraised using the Green Book approach [13]. In particular, well-being should be incorporated by using the widely collected ONS4 questions and translating the ‘life satisfaction’ question into Wellbeing-Adjusted Life Year (WELLBYs) [14], which can be easily compared across alternative policies to determine value for money though metrics such as Return on Investment (RoI) or cost-benefit analysis (CBA). To date, the only large-scale analysis of WELLBYs has come from a large-scale national evaluation of green SP, focusing on nature-based activities in the UK. Findings from this study showed statistically significant improvements in wellbeing as measured by the Office for National Statistics wellbeing measures (ONS4) across 3,339 individuals, with a 1.9-point increase in happiness, a 1.7-point increase in life satisfaction, a 1.0-point reduction in anxiety, and a 1.3-point increase in sense of worthwhileness [15]. From this, the authors estimated the social return on investment was £1.88-£2.42 per £1 invested. However, in line with previous work, its narrow focus on nature-based activities means that wellbeing effects to other referrals are uncertain.

Thus, to address the critical gap, the present study leveraged data from administrative records obtained from Access Elemental, a digital SP platform used by health and social care professionals, community development workers and other service providers to keep track of SP activities and their impact from the point of referral. While Elemental does not comprise a random sample of sites or patients in the UK, it does have good geographical coverage in England (45% of Integrated Care Boards), Wales (57% of Welsh University Health Boards) and Northern Ireland (80% of Health and Social Care Trusts), alongside smaller representation in Scotland (13% of Scottish Health and Social Care Partnerships). A challenge with the Access Elemental dataset is that it only contains records for patients who received SP, precluding the creation of non-referred counterfactuals possible in other primary care data sources. However, compared to primary care data, Access Elemental data has the major advantages of multi-stakeholder data input, allowing for examining other SP pathways in addition to the GP LW model, and richer information on details of referrals including embedded pre-post outcome measures in some sites. As such, while no dataset can provide a complete picture of SP practice, Access Elemental provides an important data source to triangulate with data from local evaluations and experimental studies. Further, to strengthen the robustness of analyses, we applied more sophisticated statistical analyses than previously used for exploration of SP, employing growth curve modelling within the Bayesian framework, which provides more flexibility for modelling pre-post longitudinal data.

Specifically, our study aimed to: (1) examine changes in wellbeing before and after SP, comparing outcomes across multiple wellbeing domains; (2) assess potential heterogeneous impacts of SP by individual characteristics; and (3) quantify wellbeing improvements in monetary terms and offer some insights into RoI. These aims are critical in building the evidence base for SP considering its global proliferation and the NHS’s pivotal moment in transforming the health and care system toward neighbourhood health.

## Materials and Methods

### Data

To date, Access Elemental has data of over 600,000 SP cases from 482,575 patients. In this study, data were restricted to patients with repeated wellbeing measures between two time points within 1-6 months between 2017-2025 in the UK, with a mean follow-up of 2.5 months (SD=1.2). For a small number of patients with more than one SP case recorded (∼2-4%), the first case was used. The analytical sample sizes ranged from 14,657 to 19,627 for different wellbeing measures, with patients from over 300 SP sites covering all countries and regions in the UK (Figure S1). See the sample selection diagrams in the Supplementary Material (Figure S2).

### Measurement

#### Wellbeing

The most commonly used wellbeing measures in the Access Elemental data were the short Warwick-Edinburgh Mental Wellbeing Scale (SWEMWBS) and ONS4. The SWEMWBS is a widely used validated tool for measuring mental wellbeing and detecting clinically meaningful changes [16,17]. It includes seven positively worded items with five response categories from ‘none of the time’ to ‘all the time’. A total score was generated by summing the scores across seven items, which was then transformed into metric scores, ranging from 7 to 35 [18].

The ONS4 questions cover happiness (affective wellbeing), anxiety (affective wellbeing), life satisfaction (evaluative wellbeing), worthwhileness (eudemonic wellbeing), each measured on an 11-point Likert scale (0-10)[19].

#### Confounders

Socio-demographic covariates included gender (female, male), age (Under 30, 30-49, 50-69, 70+), index of multiple deprivation (quintiles: 1-most deprived to 5-least deprived), and urbanicity (urban, rural). Both area deprivation and urbanicity were derived via postcode linkage to urban rural classification and index of multiple deprivation at Lower Layer Super Output Area (LSOA) in each country. We also included referral route indicating whether patients were referred from medial (e.g. GP, secondary care) or non-medical routes (e.g. education, local authority). Although ethnicity was available, as an optional measure, it had a high missing rate (>83%), so was excluded from the analysis.

#### Monetisation & ROI

As a key Wellbeing-Adjusted Life Years (WELLBY) metric, a one-point change in life satisfaction per year on the 0-10 scale is assigned a monetary value of £13,000, with a low-high range of £10,000 to £16,000 in 2019 prices based on analysis published in HM Treasury Green Book [14]. As our follow-up period was 2.5 month, we recalculated the monetary value for this time period rather than for the 12 months it typically represents, although this reflects a conservative estimate given analyses from previous studies suggest effects are maintained for longer.

To calculate ROI, we used the most recent Personal Social Services Research Unit (PSSRU) Unit Costs of Health and Social Care, which estimate the cost per person for a SP referral to be £466 [20].

### Statistical analysis

Data were analysed using Bayesian growth curve modelling. It allowed us to examine person-specific and average changes between two time points, with random intercept and slope. For the main analysis, we fitted an unconditional linear growth model to the full analytical sample for each outcome measure (SWEMWBS: N=19,627 ONS4: N=14,657). We conducted sensitivity analyses restricting to patients with recorded intervention prescriptions between the pre-post data time points (SWEMWBS: N=10,413 ONS4: N=5,677), and the subsample in 2023-2025 excluding data collected before or during the COVID-19 pandemic (SWEMWBS: N=9,276 ONS4: N=7,738). Sensitivity analyses were also performed by adjusting entropy balancing weights to address characteristic imbalance between the analytical sample and the full cohort in gender, age, area deprivation, urbanicity, country and referral route (SWEMWBS: N=15,001 ONS4: N=11,720). Further analyses were conducted to examine if changes of wellbeing differed by individual characteristics by fitting conditional growth models using data from the subsample without missing covariates (SWEMWBS: N=15,001 ONS4: N=11,720). The Bayesian growth curve models were fitted using non-informative priors, 2000 iterations, a burn-in of 1000 and a thinning of 5 and using Markov Chain Monte Carlo (MCMC) algorithms, implemented in JAGS. All analyses were conducted in in R 4.4.1.

## Results

### Demographics

The SWEMWBS and ONS4 analytical samples were broadly comparable to the overall Elemental patient cohort regarding gender, age, and area deprivation (Table 1). There were, however, some imbalances in geography and referral route. Specifically, patients from rural areas were overrepresented in the SWEMWBS sample but underrepresented in the ONS4 sample. Similarly, there was an overrepresentation of patients from Northern Ireland in the SWEMWBS sample, but an underrepresentation in the ONS4 sample. In both SWEMWBS and ONS4 samples, there was an underrepresentation of patients from non-medical route. The majority of data (>60%) were collected between 2021 and 2024.

**Table 1.** Sample characteristics of the SWEMWBS analytical sample.

| Variables | Analytical sample<br>SWEMWBS |  | Analytical sample<br>ONS4 |  | All data |  |
| --- | --- | --- | --- | --- | --- | --- |
|  | Valid N<sup>†</sup> | % | Valid N<sup>†</sup> | % | Valid N<sup>†</sup> | % |
| Gender: Female | 19,274 | 64.1% | 14,484 | 65.9% | 584,255 | 61.8% |
| Gender: Male | 19,274 | 35.9% | 14,484 | 34.1% | 584,255 | 38.2% |
| Age: 29 or under | 19,558 | 15.5% | 14,656 | 12.4% | 597,684 | 15.2% |
| Age: 30-49 | 19,558 | 31.6% | 14,656 | 30.7% | 597,684 | 31.6% |
| Age: 50-69 | 19,558 | 36.4% | 14,656 | 36.7% | 597,684 | 31.9% |
| Age: 70+ | 19,558 | 16.5% | 14,656 | 20.2% | 597,684 | 21.3% |
| IMD: 1 (most deprived) | 17,394 | 42.0% | 11,937 | 36.3% | 468,524 | 37.1% |
| IMD: 2 | 17,394 | 21.0% | 11,937 | 21.0% | 468,524 | 22.3% |
| IMD: 3 | 17,394 | 16.0% | 11,937 | 17.1% | 468,524 | 16.5% |
| IMD: 4 | 17,394 | 13.3% | 11,937 | 15.7% | 468,524 | 14.2% |
| IMD: 5 (least deprived) | 17,394 | 7.8% | 11,937 | 9.9% | 468,524 | 9.9% |
| Area: Rural | 17,394 | 19.9% | 11,937 | 8.1% | 468,524 | 12.1% |
| Area: Urban | 17,394 | 80.1% | 11,937 | 91.9% | 468,524 | 87.9% |
| Referral route: non-medical | 16,939 | 10.1% | 14,416 | 8.5% | 542,663 | 17.1% |
| Referral route: medical | 16,939 | 89.9% | 14,416 | 91.5% | 542,663 | 82.9% |
| Country: England | 19,597 | 68.3% | 14,626 | 88.0% | 600,001 | 82.5% |
| Country: Wales | 19,597 | 5.8% | 14,626 | 8.8% | 600,001 | 10.1% |
| Country: Scotland | 19,597 | 9.8% | 14,626 | 2.6% | 600,001 | 2.7% |
| Country: Northern Ireland | 19,597 | 16.1% | 14,626 | 0.6% | 600,001 | 4.7% |
| Case year:2018 or before | 19,627 | 1.8% | 14,657 | 0.0% | 601,529 | 2.0% |
| Case year:2019 | 19,627 | 7.4% | 14,657 | 0.4% | 601,529 | 3.8% |
| Case year:2020 | 19,627 | 7.3% | 14,657 | 5.3% | 601,529 | 7.5% |
| Case year:2021 | 19,627 | 16.3% | 14,657 | 15.1% | 601,529 | 14.3% |
| Case year:2022 | 19,627 | 20.0% | 14,657 | 26.4% | 601,529 | 21.1% |
| Case year:2023 | 19,627 | 21.9% | 14,657 | 24.8% | 601,529 | 22.8% |
| Case year:2024 | 19,627 | 22.8% | 14,657 | 24.9% | 601,529 | 21.4% |
| Case year:2025 | 19,627 | 2.6% | 14,657 | 3.2% | 601,529 | 7.2% |

Notes: † The unit of N for the SWEMWBS and ONS4 analytical samples is patient (only one case per patient was kept); whereas the unit of N for all data is case (one patient may have more than one social prescribing case) considering some variables (e.g. age, referral route and case year) are at case level

### Wellbeing changes

Figure 1 presents the estimated average growth trajectories and their 95% highest density intervals (HDI) from unconditional growth models (also see Table S1). On average, SWEMWBS increased by 3.31 points (95% HDI: 3.26-3.37) between two time points within 1-6 months. Happiness increased by 1.59 (95% HDI: 1.55-1.63), life satisfaction by 1.57 (95% HDI: 1.54-1.61), and worthwhile by 1.4 points (95% HDI: 1.36-1.43). And anxiety decreased by 1.45 points (95% HDI: -1.49 to -1.41). Monetisation Using monetisation methods, a 1.57-point increase in life satisfaction over a mean period of 2.5 months was equivalent to £4,252 in 2019 prices (low: £3,271, high: £5,233). If restricting to the intervention subsample, the monetised value of 1.69-point increase was £4,577 (low: £3521, high: £5,633). Based on this and the PSSRU estimates, the RoI for SP is estimated to be £4,252 / £466 = £9. Specifically, every pound invested in SP is likely to accrue £9 [low = £7, high = £11] in wellbeing benefits.

**Figure 1.**
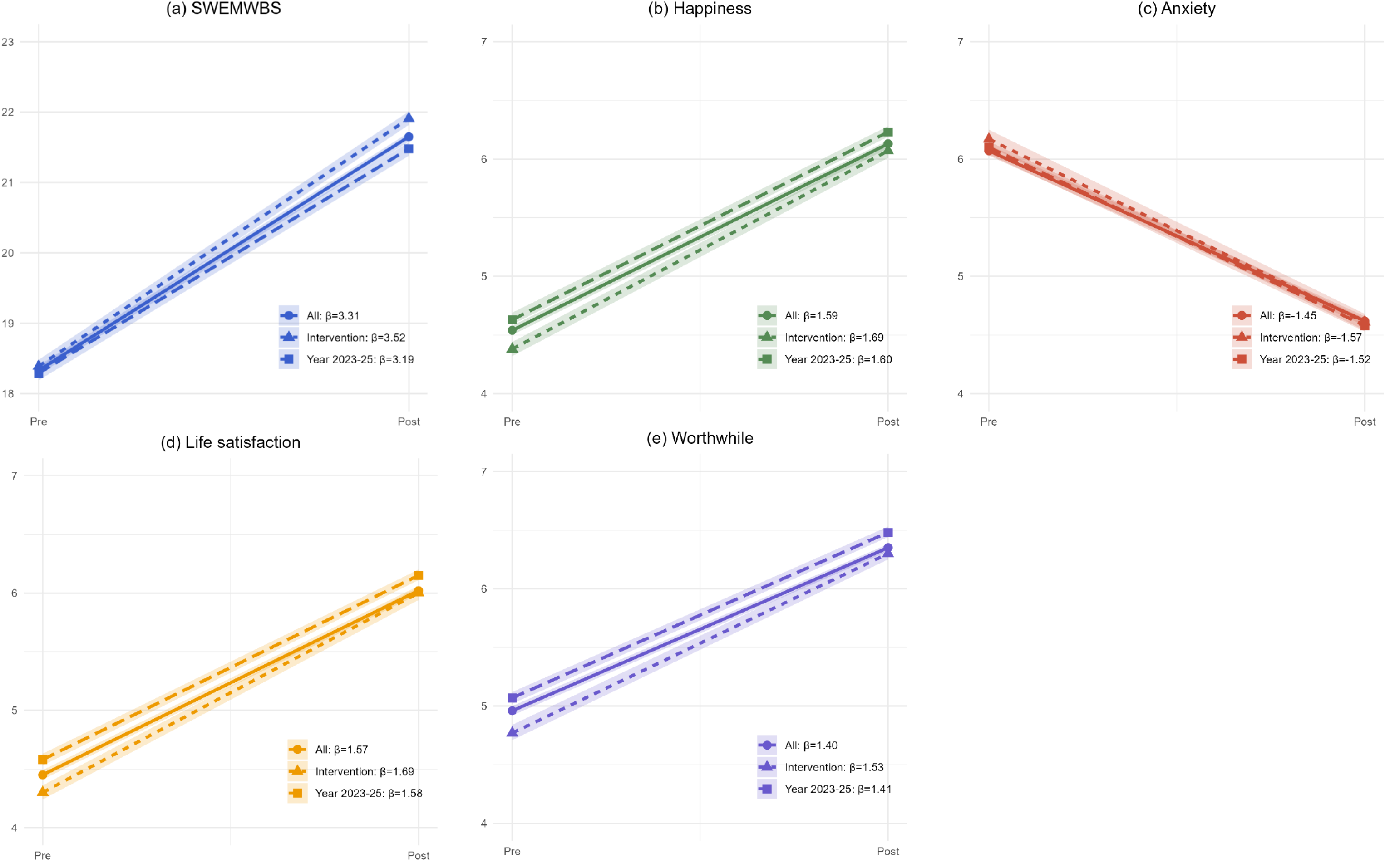
Predicted average trajectories and their 95% highest density intervals (HDI) from unconditional Bayesian growth curve models

### Sensitivity analyses

Restricting analyses to the intervention subsample yielded marginally faster rates of increase/decrease compared to the main analyses (Table S1). For example, life satisfaction was estimated to increase by 1.69 points (95% HDI: 1.64-1.75), compared to 1.57 in the main analysis. When limited to 2023–2025, change rates were similar to those in the main analyses (Table S1). Notably, however, levels of happiness, life satisfaction, and sense of worthwhile were consistently higher in the 2023–2025 subsample. Finally, sensitivity analyses adjusting entropy balancing weights yielded results consistent with the main analyses (Figure S3).

### Sub-group differences

Based on conditional growth models, while wellbeing improved across all age groups, older adults aged 70 or over had consistently slower improvement across all outcomes compared to younger adults under 30 (Figure 2). This pattern aligns with the negative intercept-slope covariance, considering older adults had higher levels of baseline wellbeing. There was some evidence that improvement rates differed by sex for happiness and anxiety, though the differences were only marginal (Figure 3). There was little evidence that wellbeing improvements differed by area deprivation, urbanicity or referral route (Table S2).

**Figure 2.**
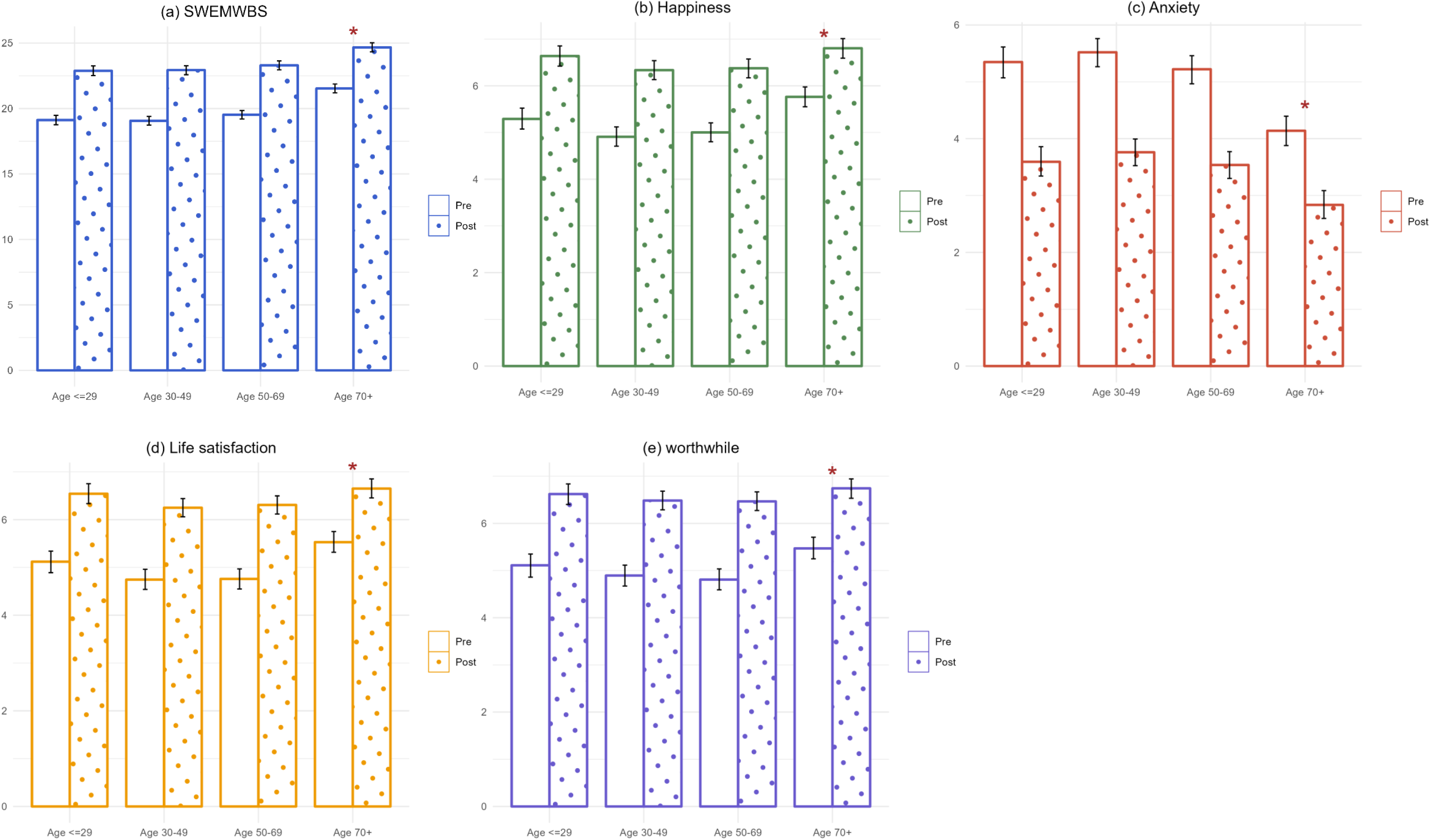
Predicted pre and post wellbeing measures and their 95% highest density intervals (HDI) by age groups based on conditional Bayesian growth curve models

**Figure 3.**
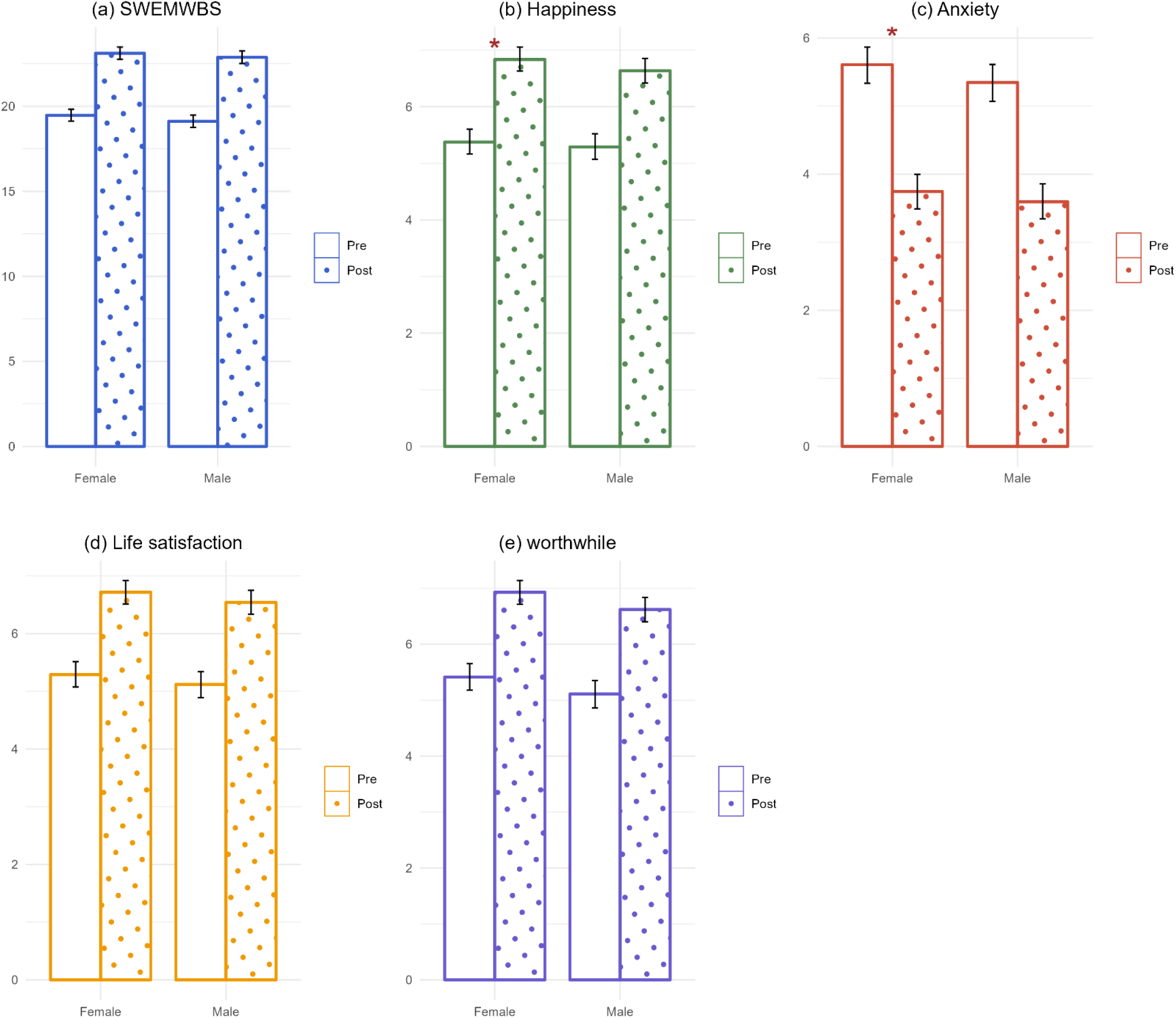
Predicted pre and post wellbeing measures and their 95% highest density intervals (HDI) by gender based on conditional Bayesian growth curve models

## Discussion

Analysing data from 14,657 to 19,627 patients from over 300 SP sites, our study provided the first large-scale national analysis of the wellbeing impacts of SP in the UK. Our findings demonstrate consistent and sizeable improvements across different wellbeing measures, including general mental wellbeing measured by SWEMWBS, affective (happiness and anxiety), evaluative (life satisfaction) and eudemonic (worthwhileness) wellbeing measured by ONS4. The increase in life satisfaction is equivalent to a conservative monetary estimate of £4,252 over a mean period of 2.5 months (2019 prices), representing a RoI of £9. We found little evidence that the wellbeing changes differ across socio-demographic groups, indicating a broad applicability of SP interventions.

Overall, our study support findings from previous work, showing a positive impact of SP on wellbeing outcomes [5,6,15]. Notably, the magnitudes of wellbeing changes in our study are largely comparable to those found in the green SP study using ONS4 measures, despite its specific focus and differential analytical methodology [15]. Regarding timeframe for wellbeing impact, our study’s mean follow-up of 2.5 months (1-6 months) aligns with previous studies using SWEMWEBS or WEMWEBS measures (1.9-4 months) [6], providing evidence for wellbeing impacts over short and intermediate durations. It remains unclear whether wellbeing improvements are sustained over longer periods, although previous trials of SP interventions suggest maintenance does occur [21]. Compared to previous work, our study expands on the existing literature by drawing on a large national dataset, which is not limited to specific geographical locations, services, or specific referral schemes. Thus, it addresses the limitations of previous work and in doing so, highlights the broad application on SP in improving wellbeing outcomes across different populations and contexts.

In relation to the health economics specifically, our findings suggest that SP yields substantial returns on investment. The 1.57-point increase in life satisfaction translate to a monetary value of £4,252 (2019 price), resulting in a £9 RoI (low = £7, high = £11). While this is a conservative estimate based solely on life satisfaction, the resulting RoI is larger than those reported in most prior small-scale studies focusing on specific interventions or patient groups [15,22,23], with the exception of one study reporting a range of £9 to £23 [24]. Our RoI calculation was based on the most recent PSSRU data for SP referral cots, but it is important to acknowledge that the underlying PSSRU data was derived from a relatively small-scale pilot study. As cost estimates of SP continue to evolve, the RoI calculation can be amended using newer values of average costs per referral. Additionally, the RoI only considers well-being as the measure of benefit. There may be other aspects of people’s lives that are positively benefited by SP that could accrue additional benefits, yet they are omitted here.

One of the major novelties of our study is analysing routine data collected in a large-scale cloud-based platform that enables tracking of diverse SP referral pathways, not just those confined to GP-LW models. However, continued efforts to improve data collection and quality are needed to fully realise their potential. Although SWEMWBS and ONS4 are the most used wellbeing measures, these wellbeing data were only recorded for one seventh patients, and fewer than 5% of patients had pre-post measures. Insight from sites suggests that the reasons for this are largely driven by whether SP sites have local-level mandates to record outcome data, meaning that the missing data can to some extent be considered ‘missing at random’ in that they are not determined by individual selection into the monitoring by patients. That said, it may be sites who are more advanced in their SP delivery are more likely to carry out pre-post outcome monitoring. The 2023 NHS England Social Prescribing Information Standard mandates that SP data is captured on patient demographics, needs and concerns, support offered (including referred to activities), and outcomes [25]. And this monitoring requirement has been reiterated in further documents, such as NHS England’s reference guide for primary care networks, giving the responsibility specifically to LWs [26]. However, it is evidently not being routinely followed, as not only were outcome data only available in some sites, but there were high rates of missingness in key measures, such as ethnicity, and time that LWs spent with patients. And in our analytical sample, only 39%-53% of patients had an intervention recorded between the pre-post time points, making it unclear whether others did not receive an intervention or it was merely unrecorded. Even when pre-post outcome measures were reportedly collected at initial and final sessions, substantial variations exist across LWs in the methods (e.g. telephone, face to face, email, home visit, office visit etc.) and timing of data collection. These ambiguities and inconsistencies significantly undermine our ability to conduct more informed analyses of how implementation factors affect outcomes. Consequently, promoting systematic outcome data collection in the future is crucial not only for service evaluation, but also for evidence-based service improvement.

Other strengths of our study include robust analytical approaches and cross-validation using multiple wellbeing measures covering different domains. However, several limitations should be noted. First, our analyses are restricted to subsamples of the Access Elemental cohort with pre-post wellbeing measures. While the analytical samples are shown to have comparable socio-demographic profiles to the whole cohort, we cannot rule out the possibly of selection bias related to unmeasured characteristics, or due to the overall representability of the Access Elemental cohort. Second, as discussed above, our study does not have any control group. A major limitation in uncontrolled pre-post design is regression to the mean, a statistical artifact that occurs when participants have extreme baseline values, leading to a natural decline when retested even without any intervention [27]. However, our analyses account for baseline-dependent growth by allowing for correlations between random intercepts and slopes and also allow us inspect person specific changes, in addition to the average rate of change, which mitigates some underlying causes of regression to the mean. The lack of control group also limits our ability to rule out confounding by temporal trends. Nevertheless, this concern is likely to be mitigated by the extended study period, with patients receiving SP at different times across almost seven years (2018-2025). Further, biases may rise from social desirability when patients are aware of the study hypotheses. However, the minimum four-week interval between assessments may reduce this bias as patients are less likely to recall their initial responses precisely.

## Conclusions

Our study significantly advances the evidence on SP impacts, demonstrating improvements across multiple wellbeing measures using national data. We find that these improvements are largely found consistently across socio-demographic groups. This indicates a broad applicability of SP in line with it being a personalised approach that focuses on what matters most to the patient, with interventions tailored to their specific needs and preferences. Based on the estimates from this study and PSSRU unit costs on SP referrals, we estimated that the RoI was £9; every pound invested in SP is associated with £9 in well-being benefits. Triangulation of these findings with other data from electronic patient health records and clinical trials will be critical to reaching a consensus on the likely impact and value for money of investment in SP services.

## Data Availability

Due to the sensitive nature of the data, the research data can not be shared publicly. Data access can be requested from the Access Elemental subject to ethical restrictions.

## Acknowledgements

This work was supported by the Economic and Social Research Council (ESRC) [UKRI1717] and the National Academy for Social Prescribing (NASP). We thank Access Elemental and all data holders for granting us the data access. LM is partially funded by the National Institute for Health and Care Research (NIHR)Applied Research Collaboration Greater Manchester (ARC-GM; NIHR200174). The views expressed in this publication are those of the author(s) and not necessarily those of the National Institute for Health and Care Research or the Department of Health and Social Care.

