## Supplement for "Assessing the impact of social prescribing on wellbeing outcomes: A national analysis of UK data"

### Supplementary Materials

Table S1. Results from unconditional Bayesian growth curve models on SWEMWBS, happiness, anxiety, life satisfaction and worthwhile

| **Main analysis: all data** | SWEMWBS (N=19,627) | | ONS: happiness (N=14,657) | | ONS: anxiety (N=14,657) | | ONS: life satisfaction (N=14,657) | | ONS: worthwhile (N=14,657) | |
|---|---|---|---|---|---|---|---|---|---|---|
| | Coef. | 95% HDI | Coef. | 95% HDI | Coef. | 95% HDI | Coef. | 95% HDI | Coef. | 95% HDI |
| **Fixed effects** | | | | | | | | | | |
| Intercept | 18.33 | [18.27, 18.40] | 4.54 | [4.50, 4.58] | 6.07 | [6.03, 6.12] | 4.45 | [4.41, 4.49] | 4.96 | [4.92, 5.00] |
| Slope | 3.31 | [3.26, 3.37] | 1.59 | [1.55, 1.63] | -1.45 | [-1.50, -1.41] | 1.57 | [1.54, 1.61] | 1.40 | [1.36, 1.43] |
| **Random effects** | | | | | | | | | | |
| SD: intercept | 4.06 | [3.72, 4.5] | 2.01 | [1.69, 2.30] | 2.26 | [1.93, 2.64] | 1.85 | [1.67, 2.07] | 2.11 | [1.86, 2.32] |
| SD: slope | 2.50 | [1.41, 3.88] | 1.49 | [0.47, 2.26] | 1.59 | [0.41, 2.59] | 1.00 | [0.25, 1.70] | 1.40 | [0.45, 1.98] |
| Covariance: intercept, slope | -0.28 | [-0.42, -0.14] | -0.53 | [-0.62, -0.45] | -0.49 | [-0.57, -0.38] | -0.53 | [-0.76, -0.44] | -0.57 | [-0.74, -0.51] |
| SD: level 1 residual | 2.15 | [1.16, 2.79] | 1.13 | [0.46, 1.62] | 1.32 | [0.40, 1.84] | 1.20 | [0.79, 1.44] | 1.07 | [0.60, 1.50] |
| **Sensitivity analysis: intervention only** | SWEMWBS (N=10,413) | | ONS: happiness (N=5,677) | | ONS: anxiety (N=5,677) | | ONS: life satisfaction (N=5,677) | | ONS: worthwhile (N=5,677) | |
| | Coef. | 95% HDI | Coef. | 95% HDI | Coef. | 95% HDI | Coef. | 95% HDI | Coef. | 95% HDI |
| **Fixed effects** | | | | | | | | | | |
| Intercept | 18.39 | [18.29, 18.48] | 4.38 | [4.32, 4.44] | 6.18 | [6.11, 6.25] | 4.30 | [4.24, 4.36] | 4.77 | [4.71, 4.84] |
| Slope | 3.52 | [3.45, 3.60] | 1.69 | [1.63, 1.76] | -1.57 | [-1.64, -1.50] | 1.69 | [1.64, 1.75] | 1.53 | [1.47, 1.59] |
| **Random effects** | | | | | | | | | | |
| SD: intercept | 4.18 | [3.77, 4.73] | 1.99 | [1.69, 2.35] | 2.15 | [1.86, 2.59] | 1.98 | [1.70, 2.26] | 2.11 | [1.87, 2.44] |
| SD: slope | 2.16 | [0.30, 3.98] | 1.42 | [0.52, 2.36] | 1.29 | [0.33, 2.54] | 1.37 | [0.46, 2.12] | 1.32 | [0.51, 2.23] |
| Covariance: intercept, slope | -0.31 | [-0.45, -0.13] | -0.53 | [-0.6, -0.43] | -0.50 | [-0.67, -0.36] | -0.55 | [-0.69, -0.47] | -0.61 | [-0.81, -0.52] |
| SD: level 1 residual | 2.23 | [0.86, 2.94] | 1.26 | [0.53, 1.71] | 1.56 | [0.71, 1.93] | 1.03 | [0.33, 1.50] | 1.19 | [0.39, 1.59] |
| **Sensitivity analysis: year 2023-2025** | SWEMWBS (N=9,276) | | ONS: happiness (N=7,738) | | ONS: anxiety (N=7,738) | | ONS: life satisfaction (N=7,738) | | ONS: worthwhile (N=7,738) | |
| | Coef. | 95% HDI | Coef. | 95% HDI | Coef. | 95% HDI | Coef. | 95% HDI | Coef. | 95% HDI |
| **Fixed effects** | | | | | | | | | | |
| Intercept | 18.29 | [18.19, 18.38] | 4.64 | [4.59, 4.69] | 6.10 | [6.04, 6.15] | 4.58 | [4.53, 4.63] | 5.07 | [5.02, 5.12] |
| Slope | 3.19 | [3.11, 3.27] | 1.60 | [1.55, 1.65] | -1.52 | [-1.57, -1.46] | 1.58 | [1.53, 1.62] | 1.41 | [1.37, 1.46] |
| **Random effects** | | | | | | | | | | |
| SD: intercept | 4.15 | [3.78, 4.63] | 1.86 | [1.65, 2.18] | 2.10 | [1.93, 2.42] | 1.96 | [1.70, 2.17] | 2.03 | [1.83, 2.29] |
| SD: slope | 2.00 | [0.27, 3.62] | 1.10 | [0.35, 1.99] | 0.88 | [0.22, 1.94] | 1.40 | [0.58, 1.98] | 1.29 | [0.60, 2.02] |
| Covariance: intercept, slope | -0.17 | [-0.41, 0.14] | -0.54 | [-0.72, -0.44] | -0.51 | [-0.8, -0.38] | -0.52 | [-0.57, -0.45] | -0.58 | [-0.71, -0.52] |
| SD: level 1 residual | 2.36 | [1.35, 2.93] | 1.27 | [0.69, 1.55] | 1.55 | [1.05, 1.75] | 0.87 | [0.30, 1.38] | 1.07 | [0.43, 1.43] |

Table S2. Results from conditional Bayesian growth curve models on SWEMWBS, happiness, anxiety, life satisfaction and worthwhile

| | SWEMWBS (N=15,001) | | ONS: happiness (N=11,720) | | ONS: anxiety (N=11,720) | | ONS: life satisfaction (N=11,720) | | ONS: worthwhile (N=11,720) | |
|---|---|---|---|---|---|---|---|---|---|---|
| | Coef. | 95% HDI | Coef. | 95% HDI | Coef. | 95% HDI | Coef. | 95% HDI | Coef. | 95% HDI |
| **Fixed effects** | | | | | | | | | | |
| Intercept | 19.12 | [18.75, 19.48] | 5.29 | [5.07, 5.52] | 5.35 | [5.07, 5.61] | 5.12 | [4.89, 5.34] | 5.11 | [4.86, 5.35] |
| Slope | 3.76 | [3.41, 4.09] | 1.35 | [1.13, 1.55] | -1.75 | [-2.00, -1.50] | 1.42 | [1.22, 1.64] | 1.51 | [1.31, 1.73] |
| Age: 30-49 (vs under 30) | -0.06 | [-0.27, 0.15] | -0.38 | [-0.53, -0.24] | 0.17 | [0.01, 0.33] | -0.37 | [-0.51, -0.24] | -0.21 | [-0.36, -0.07] |
| Age: 50-69 (vs under 30) | 0.40 | [0.20, 0.61] | -0.29 | [-0.42, -0.15] | -0.13 | [-0.29, 0.03] | -0.36 | [-0.49, -0.23] | -0.30 | [-0.44, -0.16] |
| Age: 70+ (vs under 30) | 2.40 | [2.15, 2.65] | 0.47 | [0.32, 0.63] | -1.21 | [-1.39, -1.03] | 0.41 | [0.26, 0.55] | 0.36 | [0.20, 0.51] |
| Female (vs male) | 0.35 | [0.21, 0.49] | 0.09 | [0.00, 0.17] | 0.26 | [0.16, 0.36] | 0.17 | [0.09, 0.25] | 0.30 | [0.21, 0.39] |
| IMD: 2 (vs 1) | 0.43 | [0.24, 0.61] | 0.06 | [-0.05, 0.17] | 0.06 | [-0.07, 0.19] | 0.15 | [0.04, 0.26] | 0.24 | [0.12, 0.36] |
| IMD: 3 (vs 1) | 1.17 | [0.97, 1.38] | 0.08 | [-0.05, 0.20] | 0.14 | [0.00, 0.28] | 0.17 | [0.05, 0.28] | 0.32 | [0.19, 0.44] |
| IMD: 4 (vs 1) | 1.77 | [1.54, 1.99] | 0.10 | [-0.03, 0.23] | 0.19 | [0.05, 0.34] | 0.28 | [0.16, 0.40] | 0.34 | [0.21, 0.47] |
| IMD: 5 (vs 1) | 1.33 | [1.07, 1.61] | -0.03 | [-0.18, 0.13] | 0.38 | [0.21, 0.55] | 0.11 | [-0.04, 0.25] | 0.23 | [0.08, 0.39] |
| Urban (vs rural) | -0.75 | [-0.94, -0.57] | -0.32 | [-0.47, -0.16] | 0.49 | [0.30, 0.67] | -0.22 | [-0.37, -0.06] | -0.06 | [-0.22, 0.1] |
| Medical (vs non-medical) | -1.97 | [-2.21, -1.73] | -0.45 | [-0.6, -0.31] | 0.25 | [0.08, 0.43] | -0.57 | [-0.71, -0.43] | -0.42 | [-0.58, -0.27] |
| Age: 30-49 (vs under 30) *slope | 0.12 | [-0.07, 0.31] | 0.08 | [-0.05, 0.23] | -0.01 | [-0.16, 0.15] | 0.08 | [-0.04, 0.21] | 0.08 | [-0.05, 0.21] |
| Age: 50-69 (vs under 30) *slope | 0.01 | [-0.18, 0.21] | 0.03 | [-0.11, 0.16] | 0.07 | [-0.08, 0.22] | 0.13 | [0.00, 0.25] | 0.15 | [0.02, 0.28] |
| Age: 70+ (vs under 30) *slope | -0.61 | [-0.85, -0.38] | -0.31 | [-0.45, -0.16] | 0.45 | [0.28, 0.62] | -0.30 | [-0.44, -0.17] | -0.24 | [-0.38, -0.1] |
| Female (vs male) *slope | -0.11 | [-0.25, 0.02] | 0.11 | [0.03, 0.20] | -0.11 | [-0.21, -0.01] | 0.01 | [-0.07, 0.08] | 0.01 | [-0.07, 0.08] |
| IMD: 2 (vs 1) *slope | 0.07 | [-0.10, 0.25] | 0.08 | [-0.04, 0.18] | -0.07 | [-0.19, 0.06] | 0.03 | [-0.08, 0.12] | -0.04 | [-0.15, 0.06] |
| IMD: 3 (vs 1) *slope | -0.32 | [-0.52, -0.12] | 0.05 | [-0.07, 0.17] | -0.03 | [-0.17, 0.10] | 0.06 | [-0.05, 0.17] | -0.09 | [-0.21, 0.02] |
| IMD: 4 (vs 1) *slope | -0.17 | [-0.39, 0.04] | 0.13 | [0.01, 0.25] | -0.19 | [-0.34, -0.05] | 0.03 | [-0.08, 0.14] | -0.06 | [-0.18, 0.06] |
| IMD: 5 (vs 1) *slope | 0.00 | [-0.25, 0.26] | 0.10 | [-0.05, 0.25] | -0.17 | [-0.34, 0.00] | 0.01 | [-0.12, 0.15] | -0.07 | [-0.21, 0.07] |
| Urban (vs rural) *slope | -0.19 | [-0.36, -0.01] | 0.13 | [-0.01, 0.28] | -0.02 | [-0.19, 0.16] | 0.07 | [-0.07, 0.21] | -0.09 | [-0.22, 0.06] |
| Medical (vs non-medical) *slope | -0.02 | [-0.24, 0.21] | 0.02 | [-0.12, 0.16] | 0.37 | [0.20, 0.53] | 0.06 | [-0.07, 0.19] | -0.01 | [-0.15, 0.13] |
| **Random effects** | | | | | | | | | | |
| SD: intercept | 3.55 | [3.19, 4.25] | 1.92 | [1.64, 2.29] | 2.24 | [1.93, 2.55] | 1.86 | [1.65, 2.09] | 2.05 | [1.83, 2.31] |
| SD: slope | 2.00 | [0.64, 3.99] | 1.26 | [0.28, 2.26] | 1.58 | [0.54, 2.43] | 1.16 | [0.39, 1.83] | 1.28 | [0.55, 2.04] |
| Covariance: intercept, slope | -0.11 | [-0.43, 0.29] | -0.56 | [-0.81, -0.44] | -0.48 | [-0.54, -0.4] | -0.50 | [-0.6, -0.43] | -0.57 | [-0.69, -0.5] |
| SD: level 1 residual | 2.29 | [0.62, 2.86] | 1.12 | [0.16, 1.6] | 1.29 | [0.63, 1.78] | 1.10 | [0.65, 1.42] | 1.08 | [0.46, 1.46] |

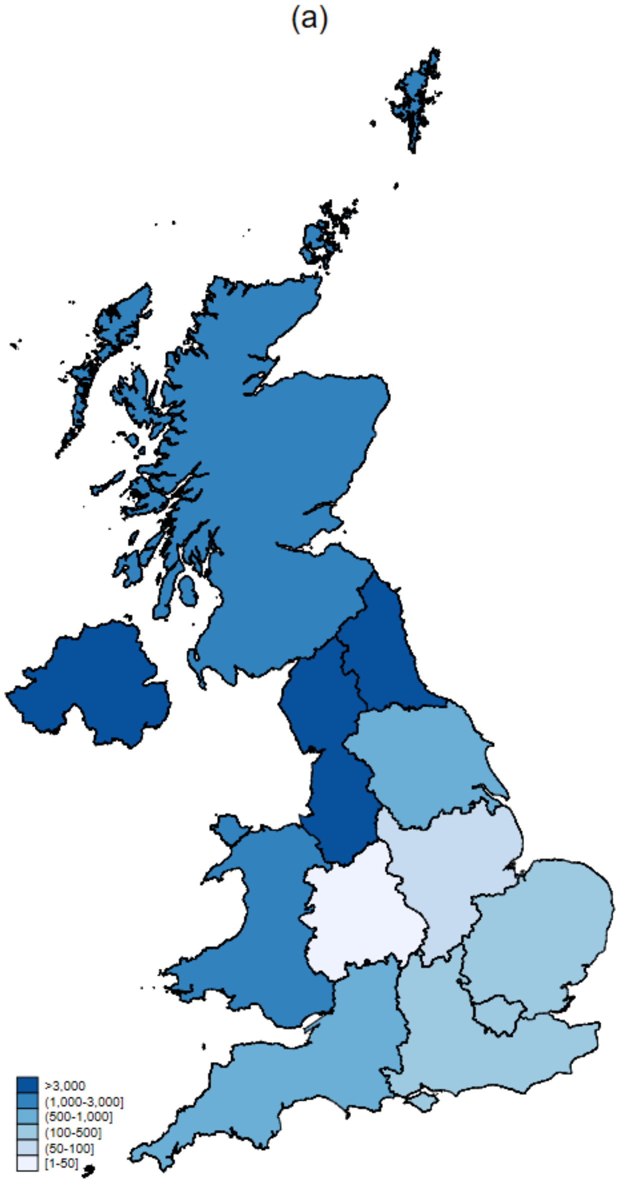


Figure S1 Sample geographic distribution across countries and regions (a) SWEMWBS (b) ONS4 (Shapefile obtained from ONS geography)

(a)

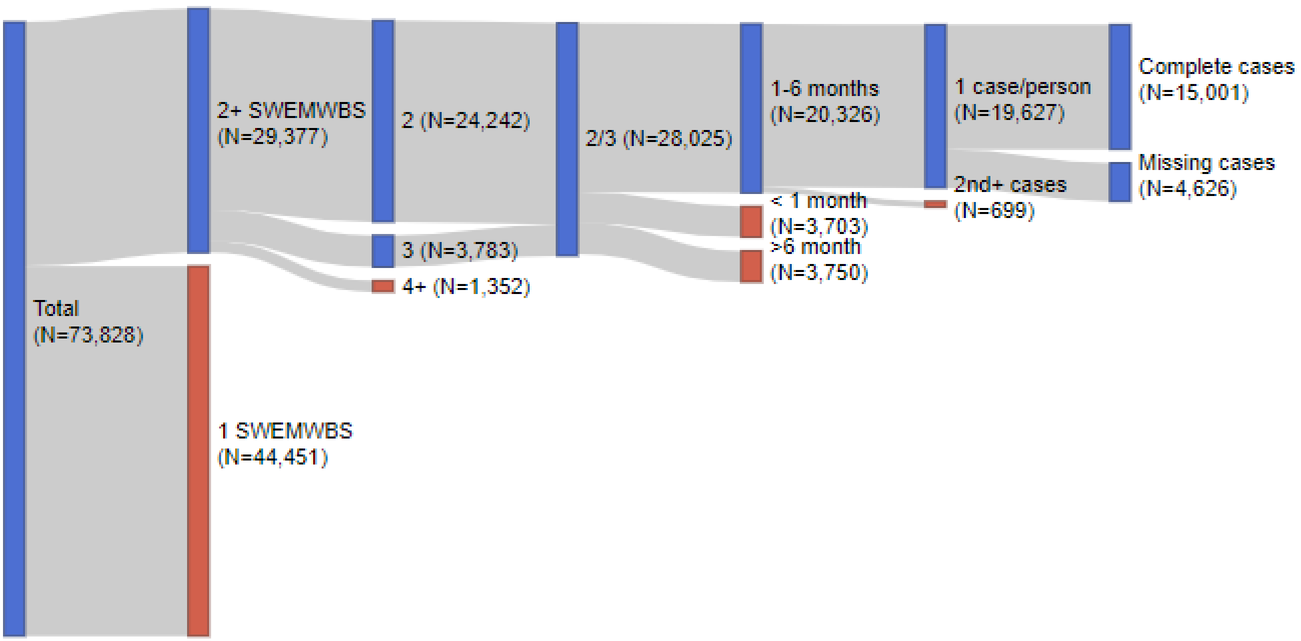


(b)

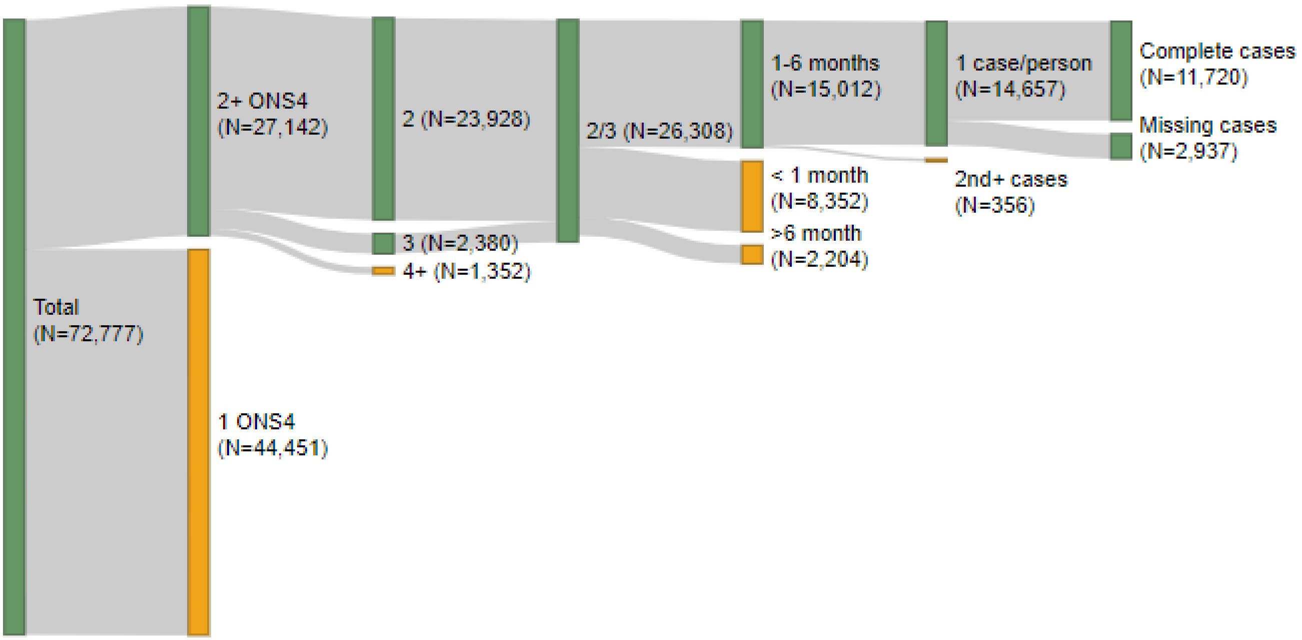


Figure S2 Sample selection diagram (a) SWEMWBS (b) ONS4

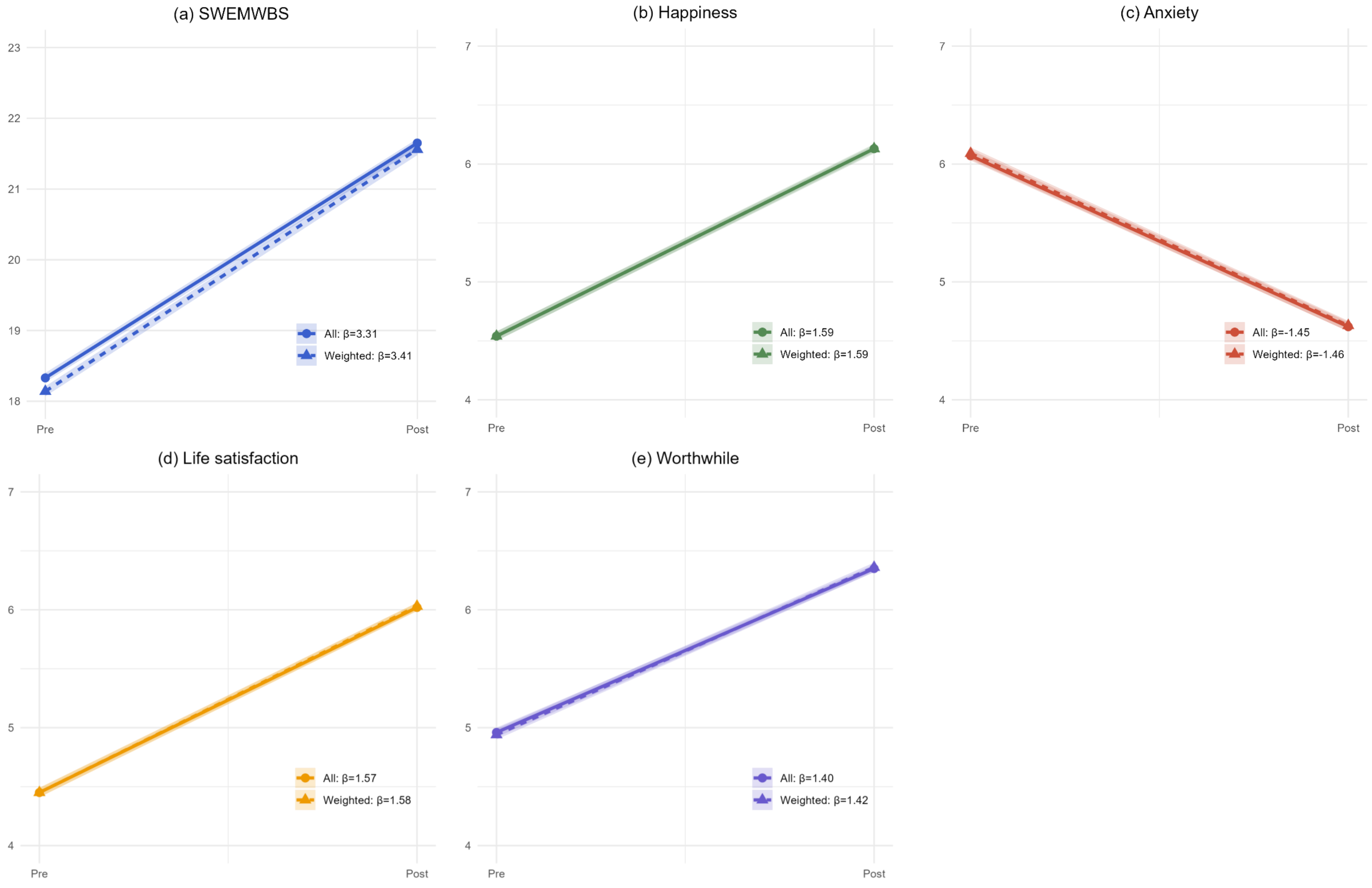


Figure S3 Predicted average trajectories and their 95% highest density intervals (HDI) from unconditional Bayesian growth curve models (weighted)
